# Baseline Physical Activity Guideline Attainment and Mortality Across Cardiovascular-Kidney-Metabolic Syndrome Stages

**DOI:** 10.64898/2026.10.28.26366776

**Authors:** Xi Luo, Yanhua Wang, Wei Zhou, Feng Yu, Junbo Sun, Kefa Xiang, Fuming Wang, Zhijun Yuan, Jiahui Bao, Shuai Zhang, Yimin Zheng, Feng Wu

## Abstract

**Background:** The 2023 American Heart Association cardiovascular-kidney-metabolic (CKM) staging framework supports risk stratification, but evidence on guideline-level physical activity across stages remains limited.

**Methods:** We emulated a target-trial protocol using NHANES 2007-2018 and National Death Index linkage through December 31, 2019. Adults aged 20 to 79 years with CKM stages 1 to 4, estimated glomerular filtration rate of at least 15 mL/min/1.73 m^2^, complete activity data, no cancer history, and eligible follow-up were included. Baseline activity was classified as meeting or not meeting the guideline. Within-stage propensity-score weights were used in weighted Cox models. All-cause and cardiovascular mortality were the primary and secondary outcomes; 5-year risks used weighted survival curves, with competing-risk cumulative incidence also estimated for cardiovascular mortality.

**Results:** Among 19,654 participants, 1,190 all-cause deaths occurred during 134,037 person-years. Baseline attainment was associated with lower all-cause mortality overall (HR, 0.774; 95% CI, 0.675-0.888). Stage-specific HRs were 1.030, 0.803, 0.642, and 0.727 for stages 1 to 4, respectively; estimates were nonmonotonic. Five-year risk differences were +0.040, -0.830, -1.790, and -3.840 percentage points; only the stage 4 interval excluded the null. Cardiovascular mortality showed a similar overall cause-specific association (HR, 0.707; 95% CI, 0.547-0.914).

**Conclusions:** Observed baseline guideline attainment was associated with mortality in this observational target-trial emulation. The findings are consistent with current physical activity recommendations but do not estimate the effect of an activity-promotion intervention; prospective trials are needed to establish clinical benefit.

**Research Perspective:** *What Is New?:* In a nationally representative US sample, observed baseline attainment of the physical activity guideline was associated with lower all-cause mortality overall, while stage-specific relative associations were nonmonotonic and five-year absolute differences were imprecise except at CKM stage 4.

*What Question Should Be Addressed Next?:* Future research should determine whether these stage-specific observational patterns are reproducible across populations and should use prospective intervention designs to investigate the mechanisms and potential clinical relevance of activity associations across CKM stages.

## Introduction

The American Heart Association’s cardiovascular-kidney-metabolic (CKM) syndrome framework organizes cardiometabolic, kidney, and cardiovascular risk across stages^1^. Physical activity is recommended for adults, including those with elevated cardiometabolic risk^2,3^. Whether associations between guideline-level activity and mortality differ across CKM stages is relevant to prevention, but stage-specific evidence remains limited.

Recent NHANES studies have examined physical activity and mortality across CKM stages. Xiao et al reported associations across stages using continuous activity measures and dose-response analyses^4^. Sartini et al examined light physical activity, with moderate-to-vigorous physical activity included as a covariate^5^. Studies in general populations also report associations between greater activity and lower mortality^6,7,8^. These studies provide relevant evidence on activity and stage-specific risk; they do not directly estimate the association of baseline attainment of the conventional activity guideline with mortality across stages using the same exposure definition.

We therefore emulated a target trial to describe the association between baseline activity guideline attainment and all-cause and cardiovascular mortality among US adults with CKM stages 1 to 4. We prespecified stage-specific estimation and five-year absolute risk differences, while interpreting estimates as observational associations rather than effects of assigned or sustained activity strategies^9,10^.

## Methods

Data, Analytic Methods, and Study Materials Availability. This study used existing, publicly available data, which are cited as data set references where they are first described below. The analysis code and derived data sets are available from the corresponding author on reasonable request; they carry no persistent identifier and are therefore not cited as a reference. No other study materials were used. This study was not preregistered: it is a retrospective analysis of existing deidentified survey data with linked mortality records, for which no preregistration platform applies. The target-trial protocol and the statistical analysis plan were specified before the analysis, are given in Table S1 in the Supplement, and are archived with the code.

### Study Design and Data Source

We specified a target trial protocol to organize eligibility, time zero, baseline activity classification, outcomes, and follow-up. The analysis compared observed baseline guideline attainment with nonattainment; participants were not assigned to an activity strategy^9,10^. The data source was the National Health and Nutrition Examination Survey (NHANES) 2007-2018,^11^ a series of cross-sectional, nationally representative surveys of the US civilian noninstitutionalized population. Mortality follow-up came from probabilistic linkage to the National Death Index^12,13^ through December 31, 2019. This study followed the STROBE^14^ and TARGET^15^ reporting guidelines. The protocol of the target trial and its emulation are given in Table S1 in the Supplement, the TARGET item reporting locations in Table S2, and a step-by-step comparison of the ideal trial and its emulation in Table S3. Because the analysis used publicly available deidentified data approved by the National Center for Health Statistics ethics review board, additional approval was not required. Participants in the original survey provided written informed consent.

### Participants

Eligibility criteria were age 20 to 79 years at the examination; CKM stage ascertainable as 1 to 4; estimated glomerular filtration rate (eGFR) of 15 mL/min/1.73 m^2^ or higher; complete physical activity data; no self-reported history of cancer; and eligibility for mortality linkage. Cancer history was an exclusion because cancer survivorship simultaneously changes activity behavior and the competing risk of death, and because cancer mortality was used as a directional test outcome. Of 59,842 examined persons, we excluded 27,454 outside the age range, 5,192 with unclassifiable stage, 5,570 at stage 0, 70 with an eGFR below 15 mL/min/1.73 m^2^, 1,867 with a cancer history, 33 ineligible for mortality follow-up, and 2 with zero follow-up time. This left 19,654 participants (Figure 1). Time zero was the examination date, at which eligibility and exposure were assessed simultaneously; participants with zero follow-up time did not enter the risk set.

**Figure 1.**
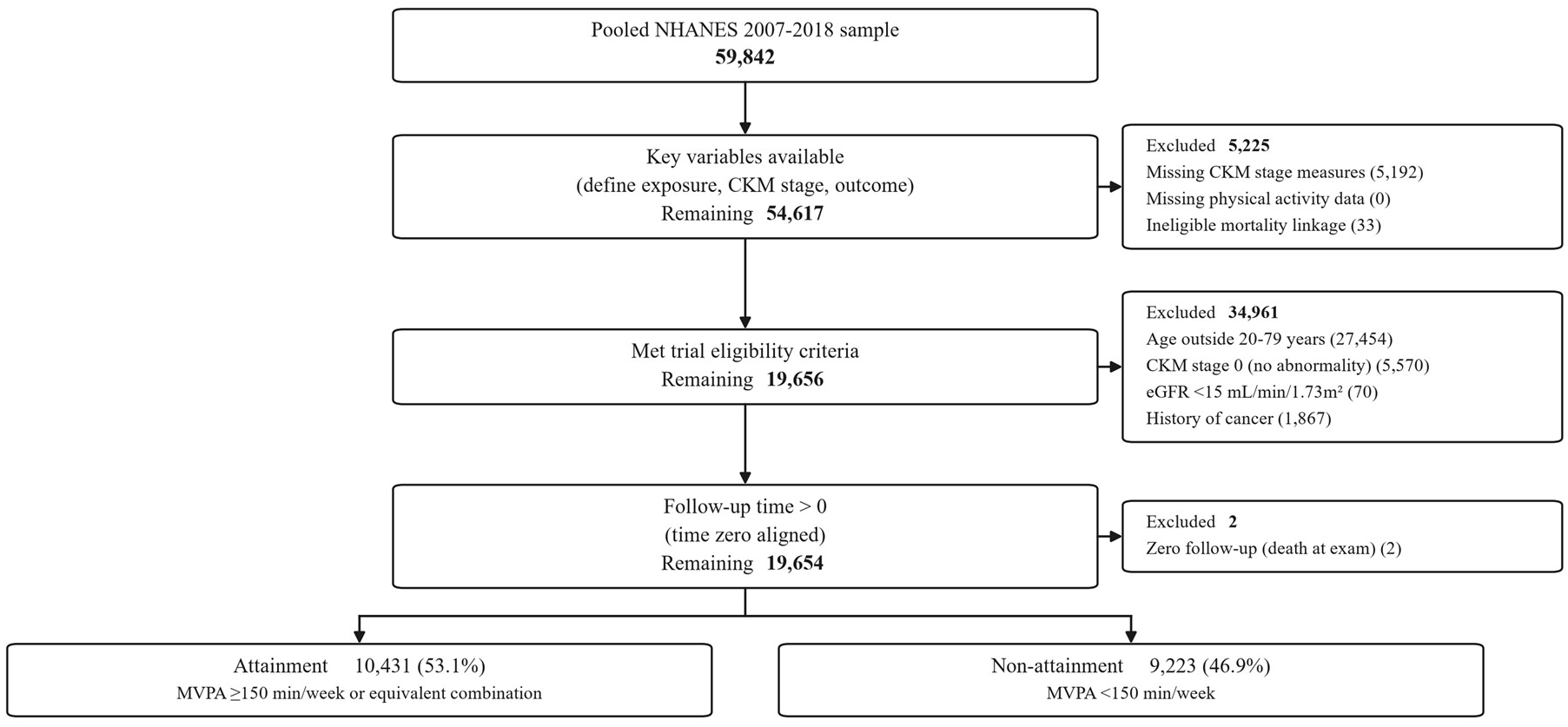
Assembly of the Analytic Cohort. Flow of participants from the 59,842 examined persons to the 19,654 participants in the analytic sample, with the number excluded at each eligibility criterion. Numbers are unweighted. CKM denotes cardiovascular-kidney-metabolic; eGFR, estimated glomerular filtration rate.

### CKM Stage

Staging followed the AHA framework^1^, with stages defined according to the data available in NHANES: stage 1, overweight (body mass index ≥25) or abdominal obesity (waist circumference ≥102 cm in men, ≥88 cm in women); stage 2, metabolic syndrome, diabetes, or chronic kidney disease; stage 3, high-risk metabolic or kidney features, used because NHANES does not ascertain subclinical cardiovascular disease (eGFR <45 mL/min/1.73 m^2^, urine albumin-creatinine ratio ≥300 mg/g, aged ≥65 years with diabetes or chronic kidney disease, or metabolic syndrome score ≥4); stage 4, established clinical cardiovascular disease, operationalized as any self-reported diagnosis of coronary heart disease, myocardial infarction, heart failure, stroke, or angina. Stage was assigned as the highest stage reached, in the order 4 > 3 > 2 > 1.

### Exposure

Exposure was self-reported moderate and vigorous activity in leisure time, work, and transportation. Meeting the guideline was defined as at least 150 minutes per week of moderate-intensity activity, at least 75 minutes per week of vigorous-intensity activity, or an equivalent combination, with vigorous minutes counted double. Activity was measured once at baseline. The comparison therefore concerns observed baseline status and does not represent sustained adherence or an intervention assigning participants to attain the guideline.

### Outcomes and Follow-up

The primary outcome was all-cause mortality. The secondary outcome was cardiovascular mortality, defined as an underlying cause of death recorded as heart disease or cerebrovascular disease. Follow-up ran from the examination date to death, loss to follow-up, or the administrative censoring date of December 31, 2019, whichever came first; no participant was lost to follow-up.

### Covariates

Baseline confounders were selected a priori from a directed acyclic graph (Figure S4 in the Supplement). They comprised age, sex, race and ethnicity, educational attainment, income-to-poverty ratio, current smoking, days of alcohol consumption, sedentary time, history of diabetes, hypertension, clinical cardiovascular disease, and chronic kidney disease, and use of antihypertensive medication and statins. Body mass index, waist circumference, blood pressure, lipids, hemoglobin A1c, eGFR, and the urine albumin-creatinine ratio were treated as potential mediators rather than confounders and were not adjusted for in the primary analysis. CKM stage was used as a stratification variable rather than a covariate. Race and ethnicity were self-reported in NHANES standard categories and entered the analysis as proxies for socially patterned exposures rather than as biological determinants. Source variables, coding, definitions, and missing rates for every variable are given in Table S4 in the Supplement.

### Statistical Analysis

Propensity scores were estimated within each CKM stage from the covariates listed above. Stabilized inverse probability of treatment weights were truncated at the 1st and 99th percentiles; weight construction and diagnostics are detailed in Supplementary Data 1 in the Supplement^16,17^. Weighted Cox proportional hazards models stratified by stage were fitted with robust standard errors. After truncation, all standardized mean differences within stage were 0.040 or less and the effective sample size was 17,437 (88.7%) (Table S5 and Figure S1 in the Supplement). Missing covariate values (income-to-poverty ratio, 9.0%; sedentary time, 0.6%) were handled by multiple imputation with chained equations (20 imputations),^18^ with propensity scores and weights re-estimated within each imputation and estimates pooled by Rubin’s rules; the fraction of missing information was 0.003 or less for every estimate.

Triglycerides and fasting glucose were ascertained only in the NHANES fasting subsample, so their missingness (51.95% and 51.50%) follows from the survey design rather than from participant behavior and is therefore plausibly missing at random; they enter only the sensitivity analysis that additionally adjusts for potential mediators and were imputed jointly with the confounders. Effect modification by stage was assessed with a joint Wald test of treatment-by-stage interaction terms.

Absolute risks were read from weighted Kaplan-Meier curves at 5 years, the primary reading window. This window falls within the observed follow-up of 16,527 participants (84.1%) from the 2007-2008 through 2015-2016 cycles, whereas the conventional 10-year window^19^ is directly supported by only 6,886 participants (35.0%) and requires extrapolation beyond administrative censoring; 10-year readings are reported in the Supplement. For cardiovascular mortality, we additionally fitted a Fine-Gray proportional subdistribution hazards model that treated noncardiovascular death as the competing event,^20^ and report the corresponding cumulative incidence function (Aalen-Johansen estimator) as a competing-risks cross-check; the absolute differences are read from weighted Kaplan-Meier curves with noncardiovascular deaths censored, and the two readings differ by no more than 0.120 percentage points over the 5-year window (Supplementary Data 2 in the Supplement). 95% confidence intervals for absolute differences were obtained by bootstrap (B = 200, with propensity scores and weights re-estimated in each replicate). Proportional hazards was assessed with Schoenfeld residuals^21^. The E value was computed for the primary estimate^22^.

Sensitivity analyses additionally adjusted for potential mediators, used untruncated weights, excluded events occurring within 2 years of baseline, used conventional multivariable Cox regression, incorporated NHANES survey weights, additionally adjusted for self-rated general health, excluded participants with mobility or any functional limitation, and re-estimated the effect with a 5-year landmark. A negative control outcome (death from unintentional injury, for which no biological pathway from activity is expected)^23^ and a directional test outcome (cancer mortality) were estimated. Continuous dose-response was modeled with restricted cubic splines (4 knots at the 5th, 35th, 65th, and 95th percentiles) with activity truncated at a plausibility ceiling of 10,080 min/wk, the total number of minutes in a week. Analyses used Python version 3.12 (lifelines, statsmodels); 2-sided P < .05 was considered statistically significant. The target-trial protocol organizes the baseline contrast but does not create randomized assignment. Because activity was observed and residual confounding cannot be excluded, the weighted contrasts are interpreted as adjusted associations rather than intervention effects. Because no family of coprimary comparisons was prespecified, no correction for multiple comparisons was applied.

## Results

### Participants

Among 19,654 participants (mean age, 48.3 years; 50.8% female; 10,431 [53.1%] met the guideline; all values unweighted), 1,190 deaths occurred during 134,037 person-years of follow-up (mean, 6.8 years; median, 6.7 years [IQR, 3.8-9.8 years]; maximum, 13.3 years). CKM stages 1 to 4 included 10,377, 4,188, 3,075, and 2,014 participants, respectively. Assembly of the analytic cohort is shown in Figure 1, and baseline characteristics by stage and guideline attainment are shown in Table 1.

**Table 1.** Baseline Characteristics by CKM Stage and Physical Activity Guideline Attainment.

| **Characteristic** | **Stage 1, attainment** | **Stage 1, nonattainment** | **Stage 2, attainment** | **Stage 2, nonattainment** | **Stage 3, attainment** | **Stage 3, nonattainment** | **Stage 4, attainment** | **Stage 4, nonattainment** |
|---|---|---|---|---|---|---|---|---|
| **No. of participants** | 6,217 | 4,160 | 2,145 | 2,043 | 1,290 | 1,785 | 779 | 1,235 |
| **Age, mean (SD), y** | 40.9 (13.9) | 45.7 (14.8) | 45.7 (13.1) | 49.9 (12.7) | 56.0 (15.3) | 61.4 (13.6) | 59.4 (12.5) | 62.4 (11.3) |
| **Female sex, %** | 43.8 | 62.8 | 45.5 | 59.7 | 42.7 | 61.1 | 34.0 | 45.1 |
| **Body mass index, mean (SD)** | 30.6 (5.3) | 31.2 (5.9) | 31.3 (7.0) | 31.9 (7.7) | 32.2 (6.8) | 32.3 (7.3) | 30.4 (6.9) | 31.0 (7.4) |
| **Current smoking, %** | 18.8 | 20.0 | 21.3 | 21.8 | 18.4 | 17.5 | 28.4 | 31.3 |
| **Diabetes, %** | 0.0 | 0.0 | 38.1 | 43.9 | 44.0 | 54.2 | 34.3 | 43.0 |
| **Hypertension, %** | 39.4 | 44.5 | 60.1 | 66.2 | 82.4 | 84.8 | 76.6 | 83.2 |
| **Clinical cardiovascular disease, %** | 0.0 | 0.0 | 0.0 | 0.0 | 0.0 | 0.0 | 100.0 | 100.0 |
| **Chronic kidney disease, %** | 0.0 | 0.0 | 32.2 | 32.5 | 40.1 | 46.1 | 25.9 | 35.5 |
| **Statin use, %** | 10.3 | 15.1 | 22.0 | 27.0 | 35.7 | 42.5 | 56.7 | 57.1 |
| **Moderate to vigorous activity, median (IQR), min/wk** | 740 (360-1,800) | 0 (0-60) | 690 (315-1,800) | 0 (0-50) | 600 (300-1,380) | 0 (0-30) | 570 (270-1,380) | 0 (0-0) |
| **Deaths, No.** | 138 | 124 | 64 | 96 | 96 | 279 | 96 | 297 |
| **Person-years, No.** | 43,099 | 29,647 | 14,280 | 13,786 | 8,440 | 12,070 | 4,925 | 7,787 |

Abbreviations: CKM, cardiovascular-kidney-metabolic; IQR, interquartile range. Values are unweighted. Stage was assigned as the highest stage reached. After weighting within stage, all standardized mean differences were 0.040 or less.

### Primary Outcome

Overall, meeting the guideline was associated with lower all-cause mortality (HR, 0.774; 95% CI, 0.675-0.888). Estimates differed by stage (P = .043 for interaction): HRs were 1.030 (95% CI, 0.779-1.361) at stage 1, 0.803 (0.565-1.142) at stage 2, 0.642 (0.491-0.840) at stage 3, and 0.727 (0.563-0.939) at stage 4 (Table 2 and Figure 2).

**Table 2.**
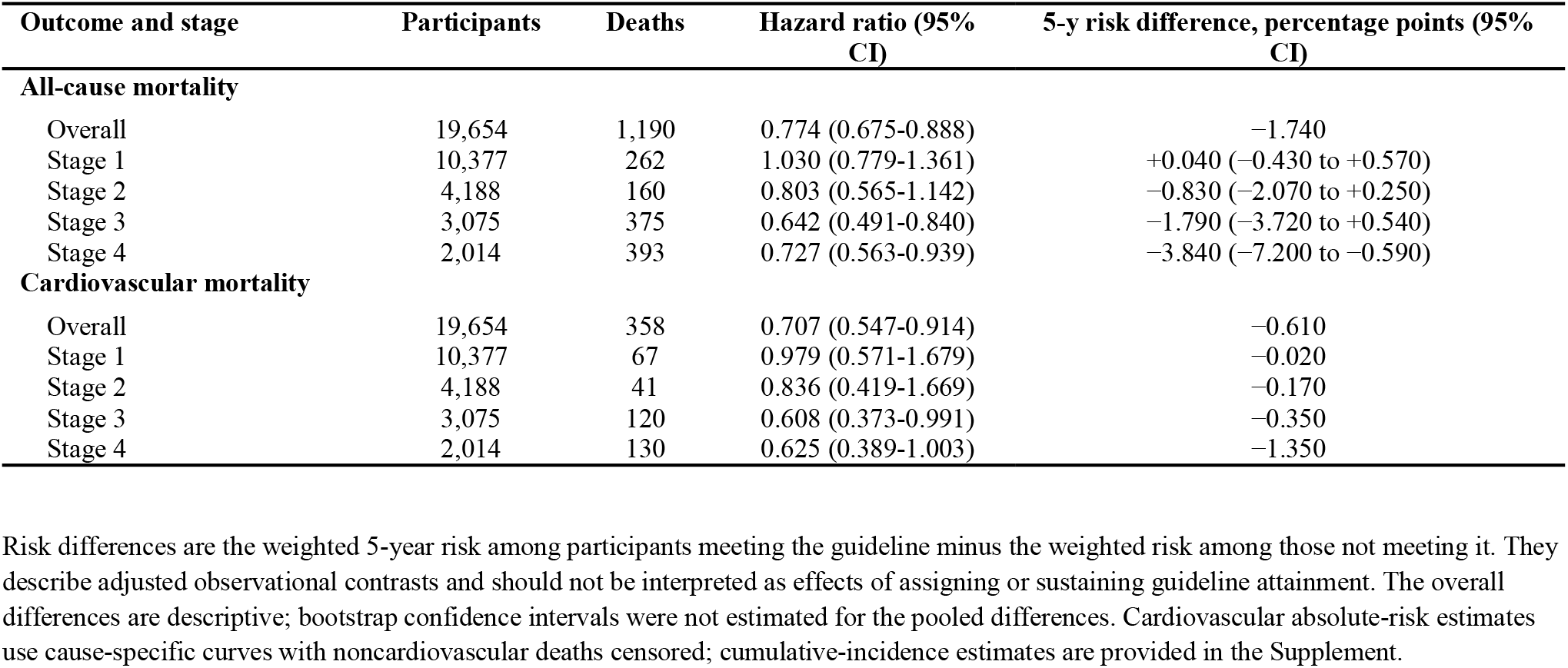
Associations Between Baseline Physical Activity Guideline Attainment and Mortality.

| Outcome and stage | Participants | Deaths | Hazard ratio (95% CI) | 5-y risk difference, percentage points (95% CI) |
|---|---|---|---|---|
| **All-cause mortality** |  |  |  |  |
| Overall | 19,654 | 1,190 | 0.774 (0.675-0.888) | −1.740 |
| Stage 1 | 10,377 | 262 | 1.030 (0.779-1.361) | +0.040 (−0.430 to +0.570) |
| Stage 2 | 4,188 | 160 | 0.803 (0.565-1.142) | −0.830 (−2.070 to +0.250) |
| Stage 3 | 3,075 | 375 | 0.642 (0.491-0.840) | −1.790 (−3.720 to +0.540) |
| Stage 4 | 2,014 | 393 | 0.727 (0.563-0.939) | −3.840 (−7.200 to −0.590) |
| **Cardiovascular mortality** |  |  |  |  |
| Overall | 19,654 | 358 | 0.707 (0.547-0.914) | −0.610 |
| Stage 1 | 10,377 | 67 | 0.979 (0.571-1.679) | −0.020 |
| Stage 2 | 4,188 | 41 | 0.836 (0.419-1.669) | −0.170 |
| Stage 3 | 3,075 | 120 | 0.608 (0.373-0.991) | −0.350 |
| Stage 4 | 2,014 | 130 | 0.625 (0.389-1.003) | −1.350 |

Risk differences are the weighted 5-year risk among participants meeting the guideline minus the weighted risk among those not meeting it. They describe adjusted observational contrasts and should not be interpreted as effects of assigning or sustaining guideline attainment. The overall differences are descriptive; bootstrap confidence intervals were not estimated for the pooled differences. Cardiovascular absolute-risk estimates use cause-specific curves with noncardiovascular deaths censored; cumulative-incidence estimates are provided in the Supplement.

**Figure 2.**
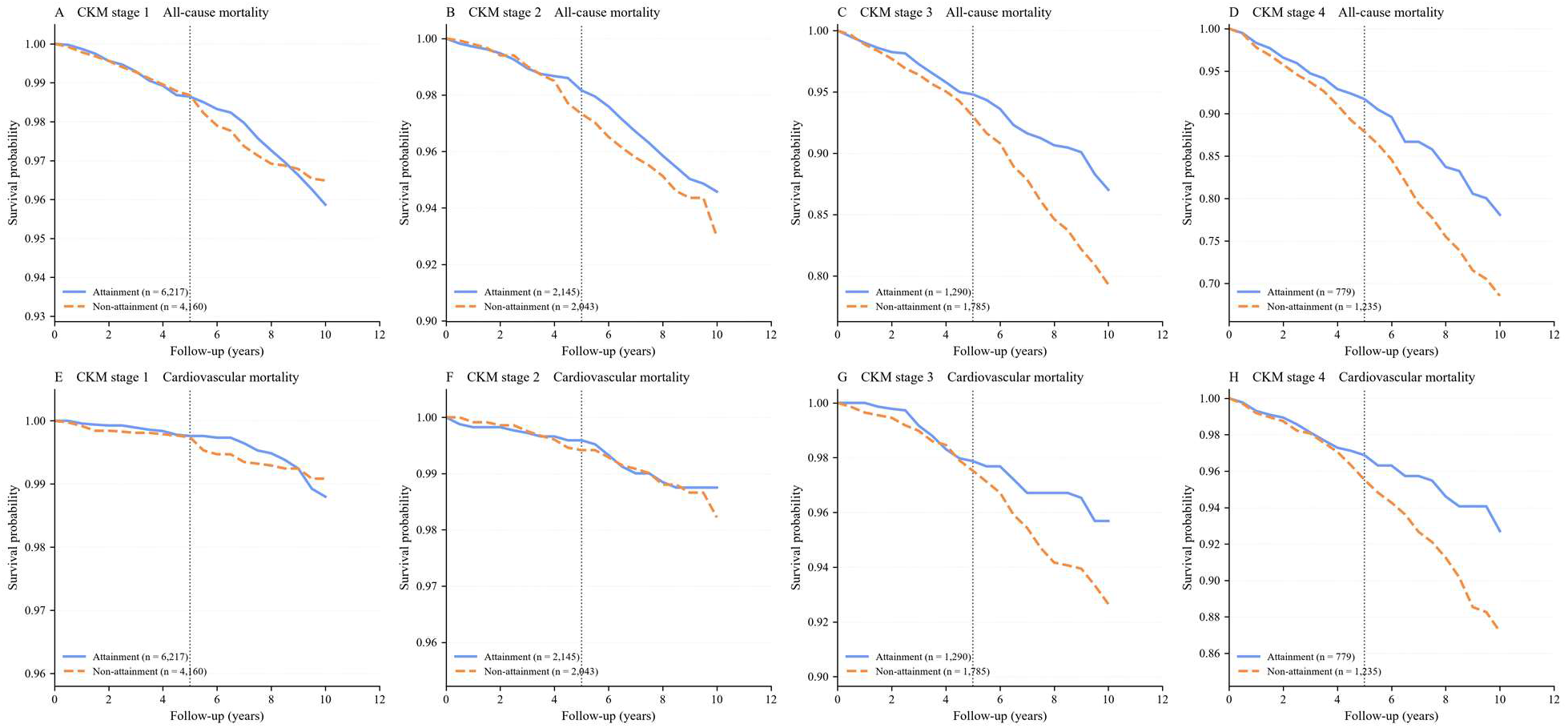
Weighted Kaplan-Meier Curves for All-Cause Mortality by CKM Stage. Curves are weighted by stabilized inverse probability of treatment weights estimated within each CKM stage. Panels A-D show all-cause mortality and panels E-H cardiovascular mortality, with noncardiovascular deaths censored. The vertical dotted line marks the 5-year reading window used for absolute risk differences in the main text.

### Absolute Differences

Weighted 5-year risks (attainment vs nonattainment) were 1.360% vs 1.320% at stage 1, 1.840% vs 2.670% at stage 2, 5.210% vs 7.000% at stage 3, and 8.290% vs 12.130% at stage 4. The corresponding risk differences were +0.040, −0.830, −1.790, and −3.840 percentage points (Figure 3). Bootstrap confidence intervals were wider on the absolute than on the relative scale. Only the stage 4 estimate excluded the null (−7.200 to −0.590 percentage points); intervals at stages 1 to 3 (−0.430 to 0.570, −2.070 to 0.250, and −3.720 to 0.540) included it. The overall 5-year risk difference was −1.740 percentage points (no bootstrap confidence interval was estimated for this pooled difference). Ten-year risk differences were larger (+0.620, −1.600, −7.730, and −9.550 percentage points) but were supported by only 35.0% of participants and required extrapolation.

**Figure 3.**
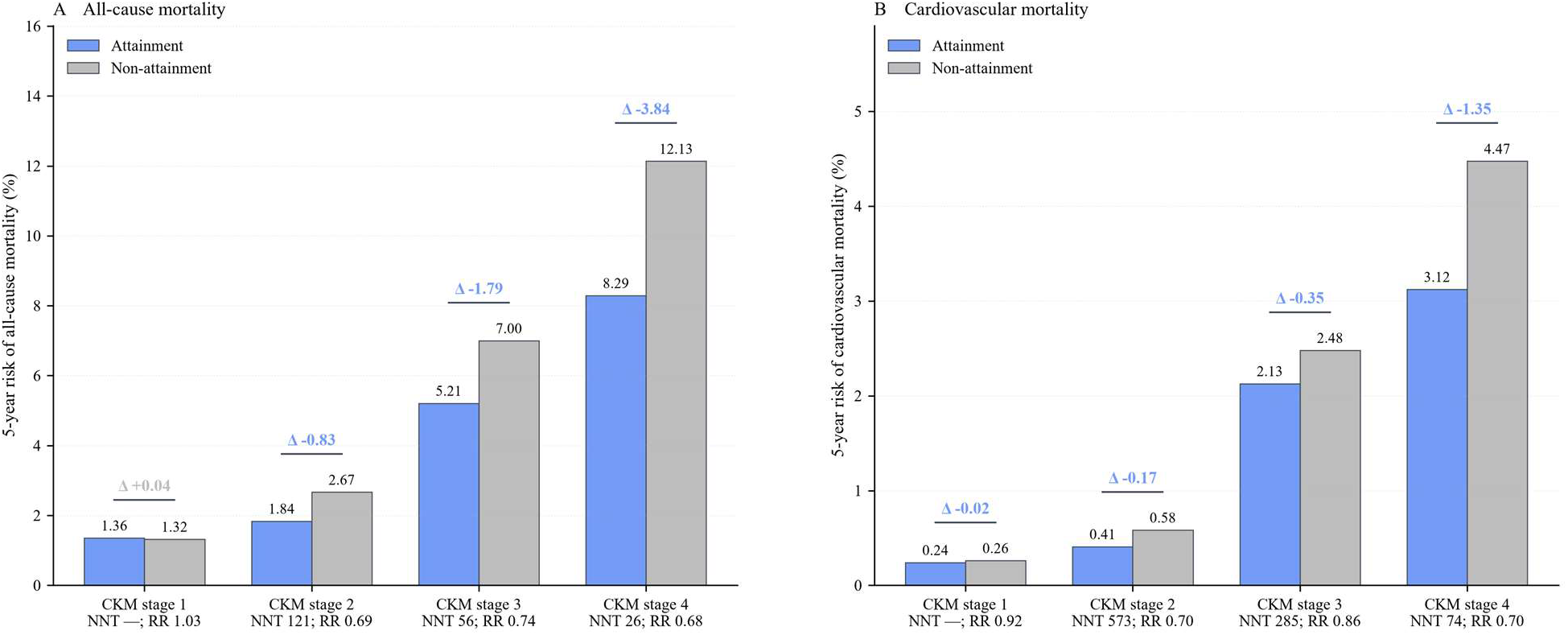
Five-Year Absolute Risk Differences by CKM Stage. Bars show the 5-year risk in the attainment vs nonattainment group, and the bracketed value above each pair is the risk difference (baseline attainment minus baseline nonattainment). Panels show all-cause mortality (A) and cardiovascular mortality (B), read from weighted Kaplan-Meier curves with noncardiovascular deaths censored. RR denotes the corresponding 5-year risk ratio. Bootstrap 95% confidence intervals (B = 200 replicates, with propensity scores and weights re-estimated in each replicate) are given in the text. These are weighted observational contrasts, not estimates of an assigned intervention.

### Secondary Outcome

Cardiovascular mortality showed a similar direction (overall HR, 0.707; 95% CI, 0.547-0.914), with stage-specific HRs of 0.979 (0.571-1.679), 0.836 (0.419-1.669), 0.608 (0.373-0.991), and 0.625 (0.389-1.003); the test for interaction did not indicate effect modification (P = .421). Five-year risk differences were −0.020, −0.170, −0.350, and −1.350 percentage points; cumulative incidence function estimates differed from cause-specific estimates by no more than 0.120 percentage points over the 5-year window (Supplementary Data 2 in the Supplement).

### Dose-Response

In spline models, hazard ratios relative to no activity were 0.828 (95% CI, 0.731-0.939) at 150 min/wk, 0.762 (0.653-0.889) at 300 min/wk, and 0.687 (0.589-0.803) at 1000 min/wk; the association was nonlinear (P = .002 for nonlinearity), with the slope steepest below the guideline threshold and flattening at higher volumes (P < .001 for association) (Figure 4). Categorical estimates referenced to inactive participants are given in Table S9 and Figure S3 in the Supplement, and the sensitivity of the spline to knot number and location and to winsorization of activity at the 99th percentile is shown in Figure S2 in the Supplement.

**Figure 4.**
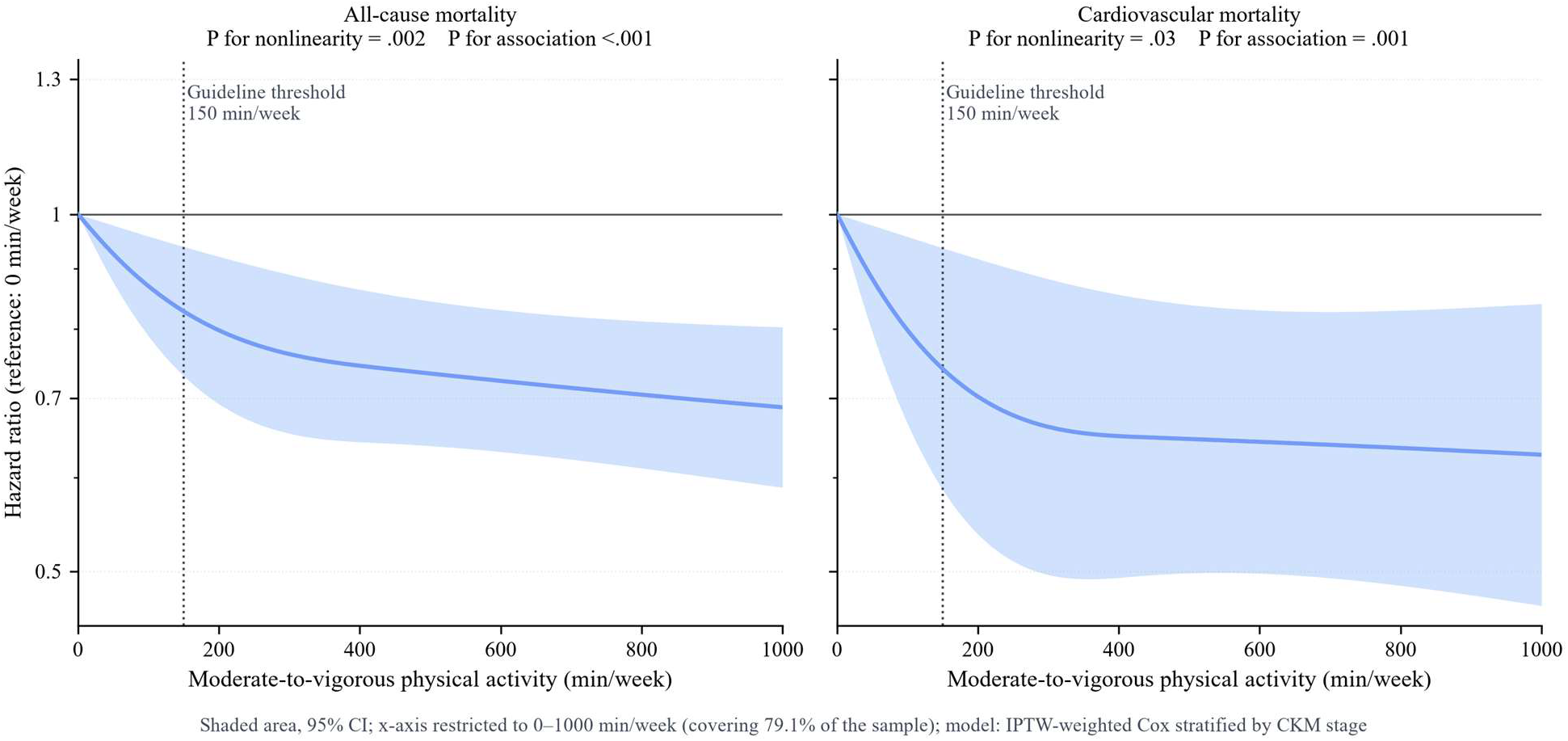
Dose-Response Association Between Moderate to Vigorous Physical Activity and All-Cause Mortality. Restricted cubic spline models with 4 knots at the 5th, 35th, 65th, and 95th percentiles, referenced to no activity, with activity truncated at a plausibility ceiling of 10,080 min/wk (the number of minutes in a week). Shaded areas indicate 95% confidence bands; the dotted vertical line marks the guideline threshold of 150 min/wk. Panel A shows all-cause mortality and panel B cardiovascular mortality.

### Sensitivity Analyses and Bias Diagnostics

The unadjusted hazard ratio was 0.443, reflecting strong confounding by age and comorbidity. Estimates were materially unchanged when potential mediators were additionally adjusted (0.796), weights were not truncated (0.783), events within 2 years were excluded (0.754), conventional Cox regression was used (0.783), survey weights were incorporated (0.802), self-rated health was additionally adjusted (0.793), and a 5-year landmark was applied (0.770; 95% CI, 0.640-0.920). Complex survey variance and weight effective sample sizes are reported in Table S8 in the Supplement, and the reverse-causation and exposure-drift analyses (A1-A3, B1-B2, and C1-C2) in Table S7 in the Supplement, where the specification checks quoted above are also tabulated with their confidence intervals (S7-3). Estimates were attenuated when participants with mobility limitation (0.880) or any functional limitation (0.916) were excluded, analyses that also removed 29% and 59% of all deaths, respectively, so the attenuation cannot be attributed to reverse causation alone. The E value for the primary estimate was 1.910 (1.500 for the confidence interval limit).

Schoenfeld residual tests did not indicate violation of proportional hazards (P = .150 for all-cause, P = .140 for cardiovascular mortality). The negative control outcome, death from unintentional injury (35 events), gave an HR of 1.955 (95% CI, 0.859-4.452), and cancer mortality gave 1.063 (0.816-1.384). The negative control estimate deviated in the direction opposite to the protective association observed for the primary outcome. With 35 events, however, its confidence interval excludes almost nothing, so this result provides too little information to calibrate residual confounding quantitatively. Alternative negative control definitions and their stage-specific estimates are given in Table S6 and Supplementary Data 3 in the Supplement.

## Discussion

### Principal Findings

In this target trial emulation of 19,654 US adults with CKM syndrome, observed baseline guideline attainment was associated with lower all-cause mortality overall. Stage-specific HRs were nonmonotonic: the estimate was above 1 at stage 1, below 1 at stages 2 to 4, and statistically significant at stages 3 and 4. The joint interaction test suggested variation across stages, but it does not establish causal effect modification. Five-year risk differences ranged from +0.040 percentage points at stage 1 to -3.840 at stage 4; only the stage 4 interval excluded the null.

### Comparison With Other Studies

Xiao et al examined physical activity and mortality across CKM stages using continuous activity measures and dose-response analyses^3^. Sartini et al studied light physical activity across stages^6^. The present analysis uses a guideline-based binary classification and a target-trial protocol to organize the observational comparison. It therefore adds a complementary exposure definition and analytic framing, not the first evidence of stage-specific activity associations or absolute risk differences. Evidence from general-population pooled and device-based studies provides additional context^6,7,8^. A target-trial emulation of physical activity and mortality in Australian women provides a related design comparison^24^.

### Interpretation

The larger absolute risk differences at later stages are compatible with their higher baseline mortality risks. However, the pattern is not a simple monotonic gradient in relative estimates, and uncertainty remains substantial: stage 3 had a statistically significant HR but its five-year risk-difference interval included the null. These results support reporting both relative associations and absolute risk differences rather than ranking stages solely by point estimates. The AHA PREVENT equations likewise emphasize absolute risk assessment across CKM health^17^.

### Limitations

Activity was measured once by self-report, and the groups were observed rather than randomized. The weighted contrasts therefore do not identify the effect of assigning individuals to attain or sustain the guideline. Residual confounding and exposure misclassification remain possible. CKM stage 3 used a study-specific operational definition based on high-risk metabolic and kidney features because NHANES did not ascertain subclinical cardiovascular disease. The stage interaction test is based on an observational model and does not by itself establish causal heterogeneity. Five-year risk differences are the primary absolute estimates; ten-year estimates require extrapolation and are reported as exploratory. The negative control had few events and its estimate was imprecise, so it cannot calibrate residual confounding.

## Conclusions

In this observational target trial emulation, baseline physical activity guideline attainment was associated with mortality across CKM stages. Stage-specific estimates were nonmonotonic, and only the stage 4 five-year risk-difference interval excluded the null. The results do not establish that assigning or sustaining guideline attainment would produce these differences. For clinical and public health practice, these estimates underscore the importance of considering absolute risk alongside relative associations: adults at CKM stages 3 and 4 had the highest baseline mortality risk, and their estimated 5-year absolute differences were larger than at stage 1. However, these observational estimates do not establish that activity-promotion interventions would reduce mortality or identify which CKM stages should receive priority for resources. Current physical activity recommendations should not be relaxed on the basis of these results. Trials or well-designed prospective emulations with repeated activity measurement are needed to estimate the effects of attaining and sustaining the guideline.

## Data Availability

NHANES 2007-2018 data are publicly available from the US Centers for Disease Control and Prevention (https://wwwn.cdc.gov/nchs/nhanes/) and the linked mortality file from the National Center for Health Statistics (https://www.cdc.gov/nchs/linked-data/mortality-files/index.html); the analysis code and derived data sets are available from the corresponding author on reasonable request.

## Acknowledgments

The authors have no nonauthor contributions to acknowledge. The authors also thank the National Center for Health Statistics for conducting NHANES and releasing the linked mortality files, and the NHANES participants. During preparation of this manuscript, the authors used an AI-assisted tool (CodeBuddy) to support coding and formatting. The authors reviewed and edited all output and take full responsibility for the content of this publication.

## Author Contributions

Feng Wu had full access to all the data in the study and takes responsibility for its integrity and the data analysis. Concept and design: Xi Luo, Yanhua Wang, Wei Zhou, and Feng Wu. Acquisition, analysis, or interpretation of data: All authors. Drafting of the manuscript: Xi Luo, Yanhua Wang, Wei Zhou. Critical revision of the manuscript for important intellectual content: All authors. Statistical analysis: Feng Yu, Junbo Sun, and Feng Wu. Obtained funding: None. Administrative, technical, or material support: Kefa Xiang, Fuming Wang, Zhijun Yuan, Jiahui Bao, Shuai Zhang, and Yimin Zheng. Supervision: Feng Wu. All authors read and approved the final manuscript. Feng Wu is the guarantor and accepts full responsibility for the overall conduct of the study and the decision to publish.

## Data Availability Statement

NHANES 2007-2018 data are publicly available from the US Centers for Disease Control and Prevention (https://wwwn.cdc.gov/nchs/nhanes/) and the linked mortality file from the National Center for Health Statistics (https://www.cdc.gov/nchs/linked-data/mortality-files/index.html); both are cited in the reference list. The analytic code is available from the corresponding author on reasonable request.

## Reporting Guidelines

This study followed the STROBE reporting guideline for observational studies and the TARGET statement for target trial emulations, both catalogued by the EQUATOR Network (https://www.equator-network.org/); completed checklists are submitted with the manuscript.

## Supplemental Material

Supplementary Data 1-3

Tables S1-S9

Figures S1-S4

No references are cited solely in the Supplemental Material.

## Non-standard Abbreviations and Acronyms

CI: confidence interval
CKM: cardiovascular-kidney-metabolic HR: hazard ratio
IQR: interquartile range
NHANES: National Health and Nutrition Examination Survey
STROBE: Strengthening the Reporting of Observational Studies in Epidemiology
TARGET: Transparent Reporting of Observational Studies Emulating a Target Trial

## References

1. Ndumele CE, Rangaswami J, Chow SL, Neeland IJ, Tuttle KR, Khan SS, Coresh J, Mathew RO, Baker-Smith CM, Carnethon M, et al. Cardiovascular-kidney-metabolic health: a presidential advisory from the American Heart Association. Circulation. 2023;148(20):1606–1635. doi:10.1161/CIR.0000000000001184.

2. Piercy KL, Troiano RP, Ballard RM, Carlson SA, Fulton JE, Galuska DA, George SM, Olson RD. The physical activity guidelines for Americans. JAMA. 2018;320(19):2020–2028. doi:10.1001/jama.2018.14854.

3. Bull FC, Al-Ansari SS, Biddle S, Borodulin K, Buman MP, Cardon G, Carty C, Chaput JP, Chastin S, Chou R, et al. World Health Organization 2020 guidelines on physical activity and sedentary behaviour. Br J Sports Med. 2020;54(24):1451–1462. doi:10.1136/bjsports-2020-102955.

4. Xiao Y, Cheng C, Zhang L, Hu J, Wang L, Shang C, Gao S, Ma B, Sun H, Zhang J. Associations of physical activity with mortality across cardiovascular-kidney-metabolic syndrome stages: evidence from NHANES 1999 to 2018. J Am Heart Assoc. 2026;15(14):e047359. doi:10.1161/JAHA.125.047359.

5. Sartini J, Rooney MR, Schrack JA, McEvoy JW, Ndumele CE, Zeger S, Selvin E, Fang M. Light physical activity and all-cause mortality in US adults across cardiovascular-kidney-metabolic syndrome stages. J Am Heart Assoc. 2026;15:e046271. doi:10.1161/JAHA.125.046271.

6. Arem H, Moore SC, Patel A, Hartge P, Berrington de Gonzalez A, Visvanathan K, Campbell PT, Freedman M, Weiderpass E, Adami HO, et al. Leisure time physical activity and mortality: a detailed pooled analysis of the dose-response relationship. JAMA Intern Med. 2015;175(6):959–967. doi:10.1001/jamainternmed.2015.0533.

7. Moore SC, Patel AV, Matthews CE, Berrington de Gonzalez A, Park Y, Katki HA, Linet MS, Weiderpass E, Visvanathan K, Helzlsouer KJ, et al. Leisure time physical activity of moderate to vigorous intensity and mortality: a large pooled cohort analysis. PLoS Med. 2012;9(11):e1001335. doi:10.1371/journal.pmed.1001335.

8. Ekelund U, Tarp J, Steene-Johannessen J, Hansen BH, Jefferis B, Fagerland MW, Whincup P, Diaz KM, Hooker SP, Chernofsky A, et al. Dose-response associations between accelerometry measured physical activity and sedentary time and all cause mortality: systematic review and harmonised meta-analysis. BMJ. 2019;366:4570. doi:10.1136/bmj.l4570.

9. Hernán MA, Robins JM. Using big data to emulate a target trial when a randomized trial is not available. Am J Epidemiol. 2016;183(8):758–764. doi:10.1093/aje/kwv254.

10. Hernán MA, Wang W, Leaf DE. Target trial emulation: a framework for causal inference from observational data. JAMA. 2022;328(24):2446. doi:10.1001/jama.2022.21383.

11. Centers for Disease Control and Prevention, National Center for Health Statistics. National Health and Nutrition Examination Survey Data, 2007-2018. US Department of Health and Human Services, Centers for Disease Control and Prevention, National Center for Health Statistics. https://wwwn.cdc.gov/nchs/nhanes/Default.aspx (2007-2018).

12. National Center for Health Statistics. The linkage of National Center for Health Statistics survey data to the National Death Index: 2019 linked mortality file (LMF): linkage methodology and analytic considerations. Published 2022. Accessed October 27, 2026. https://www.cdc.gov/nchs/data/datalinkage/2019NDI-Linkage-Methods-and-Analytic-Considerations-508.pdf.

13. National Center for Health Statistics. NCHS Public-Use Linked Mortality Files, 2019. US Department of Health and Human Services, Centers for Disease Control and Prevention, National Center for Health Statistics. https://www.cdc.gov/nchs/linked-data/mortality-files/index.html (2022).

14. von Elm E, Altman DG, Egger M, Pocock SJ, Gøtzsche PC, Vandenbroucke JP; STROBE Initiative. The Strengthening the Reporting of Observational Studies in Epidemiology (STROBE) statement: guidelines for reporting observational studies. Lancet. 2007;370(9596):1453–1457. doi:10.1016/S0140-6736(07)61602-X.

15. Cashin AG, Hansford HJ, Hernán MA, Swanson SA, Lee H, Jones MD, Dahabreh IJ, Dickerman BA, Egger M, Garcia-Albeniz X, et al. Transparent reporting of observational studies emulating a target trial: the TARGET statement. JAMA. 2025;334(12):1084. doi:10.1001/jama.2025.13350.

16. Austin PC, Stuart EA. Moving towards best practice when using inverse probability of treatment weighting (IPTW) using the propensity score to estimate causal treatment effects in observational studies. Stat Med. 2015;34(28):3661–3679. doi:10.1002/sim.6607.

17. Robins JM, Hernán MA, Brumback B. Marginal structural models and causal inference in epidemiology. Epidemiology. 2000;11(5):550–560. doi:10.1097/00001648-200009000-00011.

18. White IR, Royston P, Wood AM. Multiple imputation using chained equations: issues and guidance for practice. Stat Med. 2011;30(4):377–399. doi:10.1002/sim.4067.

19. Khan SS, Coresh J, Pencina MJ, Ndumele CE, Rangaswami J, Chow SL, Palaniappan LP, Sperling LS, Virani SS, Ho JE, et al. Novel prediction equations for absolute risk assessment of total cardiovascular disease incorporating cardiovascular-kidney-metabolic health: a scientific statement from the American Heart Association. Circulation. 2023;148(24):1982–2004. doi:10.1161/CIR.0000000000001191.

20. Fine JP, Gray RJ. A proportional hazards model for the subdistribution of a competing risk. J Am Stat Assoc. 1999;94(446):496–509. doi:10.1080/01621459.1999.10474144.

21. Grambsch PM, Therneau TM. Proportional hazards tests and diagnostics based on weighted residuals. Biometrika. 1994;81(3):515–526. doi:10.1093/biomet/81.3.515.

22. VanderWeele TJ, Ding P. Sensitivity analysis in observational research: introducing the E-value. Ann Intern Med. 2017;167(4):268–274. doi:10.7326/M16-2607.

23. Lipsitch M, Tchetgen Tchetgen E, Cohen T. Negative controls: a tool for detecting confounding and bias in observational studies. Epidemiology. 2010;21(3):383–388. doi:10.1097/EDE.0b013e3181d61eeb.

24. Nguyen B, Owen KB, Luo M, Brown W, Mielke GI, Clare PJ, Ding D. Physical activity across mid-life and mortality outcomes in Australian women: a target trial emulation using a prospective cohort. PLoS Med. 2026;23(3):e1004976. doi:10.1371/journal.pmed.1004976.

